# Improving school age nutrition and school performance through amaranth plus flaxseed food product distribution in Sidama, Ethiopia: a study protocol

**DOI:** 10.1101/2025.02.05.25321679

**Authors:** Alemselam Zebdewos Orsango, Aberash Eifa Dadhi, Mekdes Tigistu, Mehretu Belayneh Dinage, Ingunn Marie S. Engebretsen

## Abstract

Under-nutrition contributes to compromised learning, productivity and creativity in children. Primary school age children are a vulnerable group to under-nutrition, specifically anaemia and underweight. School feeding programs using locally grown foods targeting food insecure areas is one of the strategies to decrease the prevalence of under-nutrition in school children. Even if school feeding programs have been initiated in Ethiopia, the majority of vulnerable children have not benefitted from these interventions. Moreover, inadequate amount and poor quality of foods as well as sustainability of programs are challenging. Exploring underutilized and nutrient rich food sources could be one of the ways to mitigate the shortage of energy and nutrient dense food supplementation for school feeding. Amaranth is one of the few plants where leaves are eaten as a vegetable, while the seeds are used in the same way as cereals. In our previous work, we discovered that amaranth grain which grows as a wild plant has better nutrient content compared with the commonly consumed staple food maize. Also, an amaranth intervention done by this study group in younger children showed a significant effect on decreasing anaemia prevalence in children. But, we did not identify a significant weight or height change in the amaranth group. Further we found that the consumption of fish or seafood in the study area was almost null.

This study aims to assess nutritional health and to reduce under-nutrition among school children by promoting amaranth plus flaxseed food from locally grown, standardised foods in Sidama, Ethiopia.

Under this research project the following three study designs will be undertaken: a laboratory based food analysis study, a cross sectional study and an experimental pilot study.

## Introduction

Under-nutrition is a major cause of childhood morbidity and mortality in low-income countries (LICs) [1]. It is an intergenerational problem because preconception and intrauterine factors contribute to child growth and development [2, 3, 4]. Under-nutrition contributes to compromised cognitive development [5, 6, 7, 8] and affects learning, productivity and creativity [9, 10, 11]. In Ethiopia, the poor, who constitute the majority, are unable to access adequate amounts of nutrient-rich foods to meet daily dietary requirements [12, 13, 14, 15].

A synthesis of review studies from low- and middle income countries (LMICs) indicate that information about nutritional status of school aged children is not well stated [16]. In the northern part of Ethiopia a study indicated that 79.5% of school-aged children had at least one micronutrient deficiency in 2014 [17, 18]. Anaemia is one of the biggest nutritional problems in the school age going group affecting 23 % of the group [19]. In 2022, a multi-centre cross sectional study showed that the prevalence of wasting accounted for 18% of primary school aged children in Ethiopia [20]. A study from the capital city of Ethiopia indicated that 58% of school aged children had low levels of high density lipoprotein (HDL) [21].

Causes of nutritional anaemia in children are multifactorial and associated with various micronutrient deficiencies. Deficiencies of iron, vitamin B12 and folic acid are the most common causes of anaemia [22]. Despite its importance, the consumption of animal source foods and leafy vegetables in children is low in Ethiopia, [13, 23]. Further, the consumption of animal source food and leafy vegetable related with the magnitude of folic acid and Vit B12 deficiencies in school children are not explored well in Ethiopia.

School feeding in Ethiopia began in 1994, targeting food insecure areas by providing one hot meal composed of corn soya blend, vegetable oil, and salt. In 2012 Home-Grown School Feeding (HGSF) programs were introduced by the World ‘Food Program (WFP) to cover the majority of school children [24]. The HGSF is a school feeding model that provides children in schools with safe, diverse, and nutritious food sourced locally from smallholders [25]. The Ethiopian government listed School Health Nutrition (SHN) in the new Education Sector Development Plan V in 2016, and launched the SHN strategy based on the Focusing Resources on Effective School Health (FRESH) approach [26, 27, 28]. That approach aims to improve food access and educational achievement of school children through health and nutrition interventions and programmes across schools in Ethiopia [28, 29]. But, the World Food Program (WFP) report from 2013-2018 indicated the coverage of school feeding programs to be only 9% of the school children in Ethiopia [25]. In 2022 FAO evaluated the home grown school feeding program by stakeholders. The evaluation highlighted the importance of building the capacities of smallholder farmers to produce locally available, sufficient, good quality, and nutritious food to sustain the HGSF programmes [30]. Moreover, the five-year school feeding programme which is in Afar and Oromia regions in Ethiopia (2019-2024) is funded by McGovern-Dole International Food for Education and Child Nutrition Programme of the United States Department of Agriculture (USDA). This program was evaluated from 2020 to 2022 and the evaluation report indicated that the program is beneficial for food insecure households. However, sustainability is a problem as the food is imported from outside Ethiopia [31].

Regarding the effect of school feeding programs, a study from south Ethiopia found that school feeding programs decreases school absenteeism and dropout [32, 33]. The effect of school feeding on nutritional status is controversial in Ethiopia. Studies on school feeding programs found that the school feeding programs having a diverse diet has a significant effect on the nutritional status of school children [34], but a study from the rural part of Sidama region showed that the school feeding program had no effect on the anthropometric and haemoglobin measures of school children [35]. The possible reason observed under this study was that the quality of school meals provided to children was possibly not optimal to advance the nutritional status of children since the school feeding program provided a daily hot meal prepared from 150 g of dry cereals and beans in different forms [35]. Also, studies of costs and cost-efficiency of school-based feeding programs in food-insecure areas showed that fortified biscuits provided the most cost-efficient option in terms of micronutrient delivery. This study recommend that both costs, effects and sustainability should be considered carefully when designing school feeding interventions [36].

Amaranth is one of the few plants where leaves are eaten as a vegetable, while the seeds are used in the same way as cereals [37]. The seeds contain high levels of protein that is unusually complete for plant sources and are a good source of dietary fiber and minerals such as iron, zinc, copper, and manganese [38, 39, 40]. Amaranth grain has a characteristic of producing high yields even in relatively dry areas within a short period. Grain amaranth can be used as seeds or flour to make products such as cookies, cakes, pancakes, bread muffins, crackers, pasta and other bakery products. Although the grain has quality nutrients it also has a high concentration of phytic acid that can reduce the bio-availability of nutrients [41]. To solve this problem our previous study identified that homemade processing such as soaking and germination of the grain amaranth could reduce the phytate level thus increasing the bio-availability of nutrients. This finding has been supported by research from Kenya [42, 43].

In our previous work, we discovered that amaranth grain which grows as a wild plant has better iron content compared with the commonly consumed staple food maize [39]. Further, to understand the absorption of iron from the gut we did a six month intervention study on children who had anaemia. The results showed that the prevalence of anaemia decreased in the amaranth group. But, we didn’t identify a significant weight or height change in the amaranth group [38]. We believe this was due to the amount of energy content in the formulated bread. Further we found that the consumption of fish or seafood in the study area is almost null [13]. In Ethiopia, omega-3 source fatty acid foods are scarce and expensive so that the consumption of this source food is low. Flaxseed oil could be one of the alternatives to improve the intake of energy as well as the omega 3 fatty acid. Flaxseed is one of the oil plants that grows widely and its contribution to supplement omega-3 fatty acid is not well recognized in Ethiopia.

Exploring underutilized and nutrient rich food sources could be one of the ways to mitigate the shortage of energy and nutrient dense food supplementation for school feeding.

The overarching aim is to assess nutritional health among school children and to test an amaranth plus flaxseed food product provision on under-nutrition among school children in Sidama, Ethiopia. A pilot randomized trial with allocation ratio of 1:1 school.

## Methods

### Study setting and designs

The study will be conducted at Shebedino woreda rural public primary schools in Sidama Regional State, Ethiopia. The Sidama region has 36 districts (6 urban districts and 30 rural districts) including Hawassa city administration. The total population of the region is 4.4 million. Shebedino is one of the districts (woreda) and has a rural and urban wing; the rural wing of the woreda has 26 kebeles. There are 35 governmental schools in the rural wing of the woreda: from these 33 of them are primary and 2 are secondary schools. The woreda’s rural wing is divided into 26 kebeles. 33% of the woreda is classified as dega (highlands), and the remaining 67% is woyinadega (midlands), and rainfall is usually abundant at 900–1100 mm per year as the long-term mean. Inset (Kocho) and maize are the people’s main foods. Coffee, inset, fruits, avocado, mango, papaya, banana, vegetables, and chat are the most prevalent income crops in this area. Shebedino woreda has one hospital, six health centres, twenty-three health posts, and a total of 401 health professionals, including 76 health extension workers.

The three types of designs will be employed comprising fourwork-packages (WPs) each having a unique study design: i. a laboratory based experimental study design, ii. a cross sectional study design, and iii and iV an experimental pilot study.

The objectives of each of the three work packages (WPs) are outlined below, followed by a description of each

- To formulate an amaranth plus flaxseed based energy dense and micronutrient rich food product for school children 7-10 years of age in rural schools of Sidama, Ethiopia and conduct acceptability testing (WP 1).
- To assess the prevalence of and factors associated with under-nutrition in school children 7-10 years of age in rural schools of Sidama, Ethiopia (WP 2), and
- To assess the effect of an amaranth plus flaxseed based energy dense and micronutrient rich food product distribution on nutritional statusin school children 7-10 years of age in rural schools of Sidama, Ethiopia (WP 3).
- To investigate the impacts of amaranth plus flaxseed food products on the education outcomes of school-age children 7-10 years of age and its cost-effectiveness in Sidama, Ethiopia (WP4).

### Work package 1

In this WP, amaranth, flaxseed, chickpea, and emir wheat will be used for the formulation of an energy and nutrient dense recipe for the experimental school feeding pilot. Home level processing will be applied to the amaranth grain to reduce the phytate level. Amaranth grain will be soaked in water by adding 5 ml of lemon juice per 100 ml of water for 24 hours and germinated for 72 hours. After sun drying, it will be roasted and milled with a local electrical mill [37]. The chickpeas will also be soaked for 24 hours, dried, roasted, and milled. The flaxseed, emmer wheat, oats, and maize will be sorted, washed, dried, roasted, and milled.

#### Flour blend formulation

The flour blends from prepared flours will be formulated for the preparation of a recipe for standardised food preparation. The grains ratio will vary in order to assess the combination which has highest nutritive value (nutrients and energy), lowest anti-nutrient ratio to nutrient and acceptable taste, texture and preparation qualities.

#### Acceptability

A consumer acceptability test will be done at laboratory and community level by using nine-point hedonic tests. Community acceptability tests will be done with school children.

#### Nutrient analysis

The nutrients composition including minerals (iron, zinc and copper), heavy metals, amino acids, and fatty acids, and dry matter will be assessed by Institute of Marian Research (IMR) accredited by Norwegian Accreditation with registration number TEST 050. Carbohydrate content will determined by estimating difference by calculating crude protein +crude fiber +total ash +crude fat (Carbohydrate%= 100 - (fat %+protein % + ash % + moisture %). The total energy content (calories) of the food will be calculated using its fat, carbohydrate, and protein content. We’ll use standard conversion factors (Atwater factors) to do this: 4 calories per gram for protein and carbohydrates, and 9 calories per gram for fat. The formula we will use is: calories per 100 grams = (4 *carbohydrates + 4 *proteins + 9 * fats). To ensure the food is safe, we’ll test for harmful microorganisms like E. coli, Coliform bacteria, and total bacteria count using standard laboratory methods at Hawassa University food and nutrition laboratory centre. Once we confirm that the food meets safety and palatability standards according to FAO and WHO guidelines for contaminants and toxins, the intervention will be determine [44, 45].

### Work package 2

The cross-sectional survey will investigate the nutritional status among 7-10 year old primary school going children and estimate the prevalence of undernutrition and associated factors. The study will include grade 1-2 actively learning school children (7-10 years old) with parents or guardian at selected primary schools. Those who are seriously ill or have chronic illness, have severe malnutrition, have a history of any known allergy, participating in other trials, are unable to take solid food, and grade 1 or 2 children aged >10 years will be excluded from the cross-sectional survey.

**Sample Size calculation WP 2 3 and 4**

Sample size was calculated with two steps for the cross-sectional survey part (WP 2) and the experimental part (WP 3).

Sample size for the cross-sectional survey was computed using Epi Info 7.2.5.0 statistical software by considering a single population proportion with the assumptions of an alpha value of 5%, a confidence interval (CI) of 95%, a margin of error of 5%, for each outcome variable and the largest sample size was taken. Sample size ***n* = [DEFF*Np (1-p)]/ [(d**^**2**^**/Z**^**2**^_**1-**α**/2**_ **\*(N-1) + p*(1-p)]**. We calculated the sample size for each outcome variable, and ultimately chose the largest sample size needed, which was 374. This was determined based on the factor associated with undernutrition, specifically the prevalence of low high-density lipoprotein (HDL), estimated at 58.4%.

The previously reported value of the intra-cluster correlation coefficient (ICC) factor from the study reported on summarized minimum intra-cluster correlation coefficient (ICC) estimates for pupil health outcomes from school based cluster randomized trials (CRTs) across the world region was 0.01 (the minimum value) [46]. To account for the design effect, the sample size was multiplied by 1.59. The design effect was calculated with the assumption of an equal size of clusters. Using an intra-cluster correlation coefficient value of 0.01, (DIEF = 1+ [(n-1) ICC]), where ‘n’ is the average class size, which is 60 students per class, accordingly, the adjusted sample size was 595. Accounting for 10% non-response rate, the final sample size estimated was 654 participants from 36 classes in 6 schools.

The sample size for WP 3 was determined using G power statistical software. As there are no prior estimate of the outcome variables (serum level of ferritin, folate and VitB12, haemoglobin, weight, height and HDL) on grade one and two school children, the sample size was determined under the assumption of a statistical test (mean difference between two group), 95% confidence level, 80% power, conventional medium effect size of 0.5, a 1:1 ratio of intervention and control group and with ten total number of predictor variables. Accordingly, the estimated sample size became 128 (64 for each group). Then the sample size was adjusted considering a design effect of 2 with a 10% attrition rate. The final total sample size was 282 (141 for intervention and 284 for control group).

### Sampling procedure for WP 2 and 3

Multi-stage stratified sampling technique will be used to select study participants. Six rural primary schools will be selected randomly for WP2 and individuals selected proportionally to the class size. After the cross-sectional study (WP 2) two primary schools with comparable characteristics will be selected from the six schools for WP 3. In this way the pilot study will mimic a cluster randomized trial, where the intervention is the only difference between the participants. The school getting the intervention or control intervention will be allocated randomly for the experimental and control group. The food will be fed for the whole class, and the children who participated in the cross-sectional study and their caregivers will be invited to participate in the trial pilot. Figure 1 illustrates the planned trial profile.

**Fig 1.**
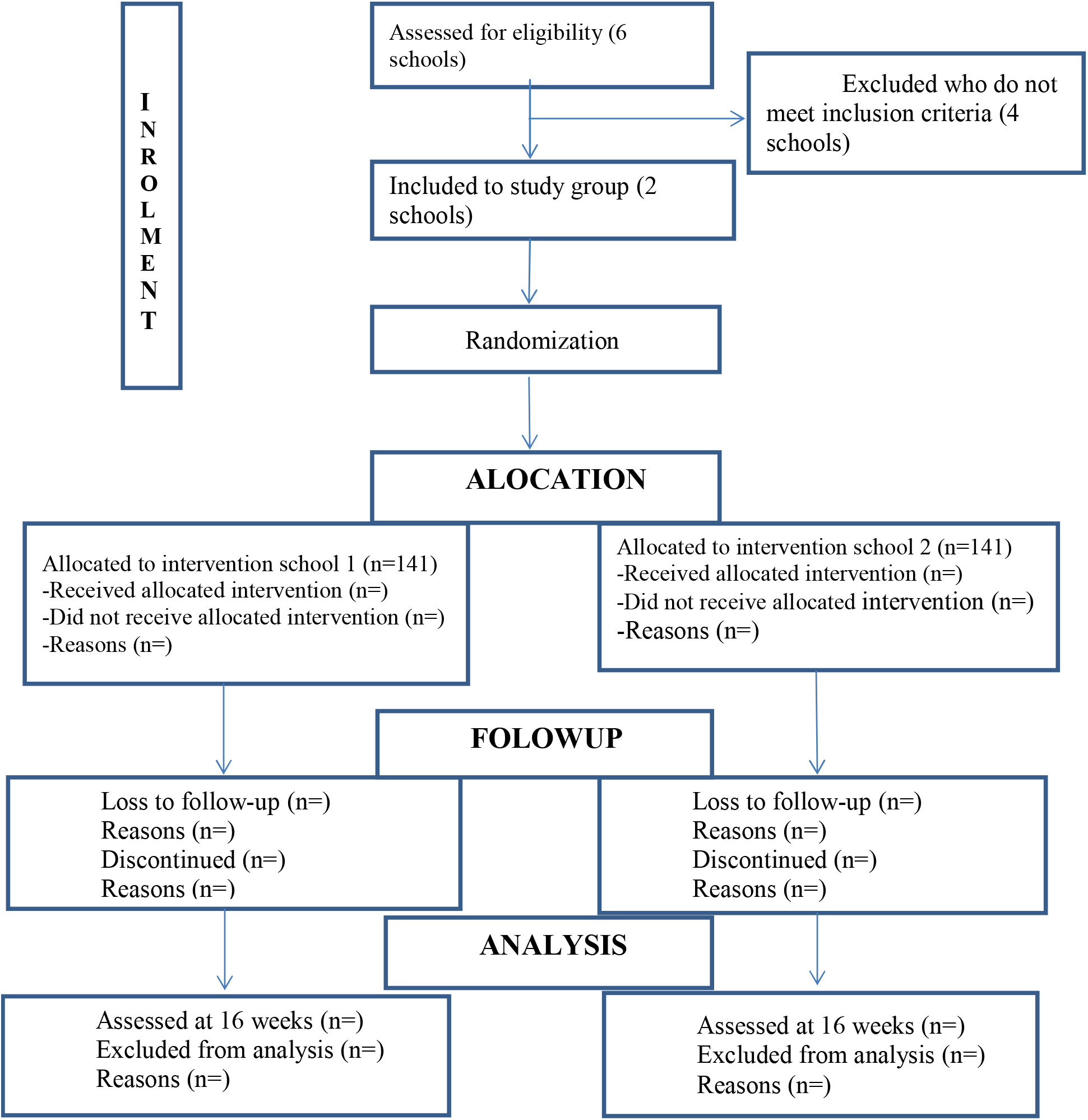
Proposed trial profile.

### Work package III and IV

The pilot experiment is described as a randomized trial with the following characteristics:

### PICO

**P** (Population): School children (7-10 years) of age in Sidama, Ethiopia

**I** (Intervention): Feeding of 200g nutrient and energy dense amaranth plus flaxseed bread five days/week for four months

**C** (Comparison): Commonly used maize bread of similar amount and colour as bread given to the intervention group

**O** (Outcome): Nutritional anemia (Ferritin, Iron, Hemoglobin, folate, vitamin B-12), HDL growth (HAZ, BMI/age-change, WAZ) and education outcome (test score and drop out) **T** (Time): 3 months

#### Eligibility

Inclusion Criteria: Accenting grade 1-2 actively learning school children (7-10 years old) with consenting parents or guardian at selected primary schools will be included.

Exclusion Criteria: Those who are seriously ill, children with severe malnutrition, children with history of any known allergy or participating in other trials, children who are unable to take solid food, grade 1 or 2 children aged >10 years, and children with chronic illness like diabetes mellitus will be excluded from the pilot study.

#### Intervention and control food

The baseline data collection and meal preparation will take place at least two months before the anticipated start of recruiting trial participants, which is tentatively scheduled for September 20, 2024. The trial will then continue for four months, with the planned end date for follow-up being January 20, 2025.

The school will be assigned randomly by external body by using lottery method to thetrial groups: i. amaranth plus flaxseed food, versus ii. maize 100% food and no intervention group. After allocation the children will be given a card that contains their identification photo and attendance sheet. The trained feeding assistance personnel will deliver the food daily to the children at school during break time. They are also responsible for monitoring the children’s health for any unusual signs and measuring the amount of food leftover. The attendance sheet will have the place of signature for feeding assistance and the sign of leftover proportion to thick every day and at the end of the week the check list will be collected from the participant and evaluated by coordinators. The correct food will be provided five days per week for 4 month in each group.

### Blinding

The trial will be double blinded, this means the distributor will be kept uninformed about the content of the food he or she is distributing and the data collectors will be kept uninformed about the trial allocation which is by school. The coordinators will distribute the food separately according to the trial allocation.

The food will be prepared based on the recommended dietary allowance (RDA), and according to RDA the target daily intake of food will be 200g for all children, considered that amount to be comfortably consumed in one session. 200 g of the proposed food will be prepared according to the description in WP 1, with an assumed proportion close to 15% flaxseed+20% amaranth+40% wheat + 20% chickpea also as an additive 20ml of sunflower oil and 10g sugar.

Both amaranth plus flaxseed food and 100% maize food will be prepared and packed separately. After packing each package will be labelled with a pre-printed sticker containing the schools’ names, in this way we will ensure the distributor gets the right food for the right participant.

#### Data collection in WP 2 and 3

Data collectors will receive training about the objectives of the study, data collection systems, interview techniques, micronutrient and anthropometric measurements, feeding procedures and field procedures prior to the data collection standardization of micronutrient and anthropometric measures will be maintained.

#### Pre-test

Pretesting of the questionnaire and food will be conducted in places away from where the actual study will be conducted. Pre-test will be conducted on 5% of the total sample size to check the tools. Materials for blood collection, anthropometry measures will be checked and prepared prior to start the actual work.

#### Interview

Using a pre-tested structured questionnaire, information on the socio-demographic (age, sex, educational status, level of income), diseases history (diarrhea, fever, vomiting, or breathing difficulties) with in the last fifteen days and any history of chronic illness repeated attack of malaria and recent diagnosed illness under medical treatment. Also 24 hours dietary history of children will be gathered through face-to-face interviews with the children’s parents.

#### Physical examination

Physical examination includes eye, mouth, neck, lymph nodes, skin, abdomen for enlargement of liver and spleen, extremities, pulse rate, respiratory rate and temperature will be assessed by trained clinical health officers.

#### Anthropometry

An electronic digital flat scale (Seca 874 weighting scale, made in Germany) will be used to measure body weight and height. Height will be taken using a Seca213 height board (Seca 213; Seca GmbH, Hamburg, Germany). Weight will be taken using Seca 874 electronic flat scale (Seca 874; Seca GmbH, Hamburg, Germany) with the child barefoot and wearing light clothing. BMI-for-age will be used to measure obesity, overweight and thinness/wasting [47].

#### Stool test

Stool samples will be collected in clean, leak-proof stool cups using conventional methods. It will be tested shortly after collection using direct procedures. Direct microscopy to identify motile protozoa and Kato-Katz stool test for the presence and severity of infection will be assessed for Ancyclostoma duodenale (hookworm), Ascaris lumbricoides (roundworm), and Trichuris trichiura (whipworm) ova.

#### Blood collection and lab analysis

A 5-6 ml of venous blood will be collected via venepuncture and then four mililitter will be transferred to serum separator tube (SST) for biochemical analysis and the remaining two will be drawn in to tube containing ethylene diamine tetraacetic acid (EDTA) for complete blood count (CBC) analysis. During blood collection all infection prevention standards will be applied. The blood samples in the EDTA tubes will be wrapped in aluminium foil, continuously shielded from light during transportation to Hawassa Referral Hospital for CBC analysis.

The collected blood in SST will be allowed to clot for 30 minutes and centrifuged at 4000rpm for 10 minutes. After centrifuging, serum will be kept in cryovial tube with the participant’s identity and will transport to Hawassa Referral Hospital using cold chain carrier at - 20^°^C. Biochemical analysis will be held at Hawassa Referral Hospital laboratory department using Cobas 6000 immediately (if possible) or stored at -70 o C until analysis.

Haemoglobin will be determined using complete blood count (CBC) machine (Sysmex Kx-21 automated analyser, Sysmex Corporation, Kobe, Japan) with a venous blood sample. Serum ferritin, iron, Vitamin B12, and folate, will be analysed by electrochemiluminescence immunoassay “ECLIA” on Cobas 6000 (e601 module). The measuring principle for CRP and _α_ 1 **-**Acid glycoprotein (AGP), will be analysed particle enhanced immunoturbidimetric assay and measured on Cobas 6000 (c501 module). Results will determined via a calibration curve which is an instrument specifically generated by 2-point calibration and a master curve provided via the reagent barcode.

### Outcome measures variables

The study will report on: WP1: nutrient content; sensory attribute of formulated food; WP2: prevalence and causes of nutritional anaemia (serum iron, haemoglobin, serum ferritin, vitB12, folic acid); level of high density lipoprotein (HDL); and WP3: change in nutritional anaemia (serum iron, ferritin, folate and Vit B-12), change in HAZ, BMI/age, WAZ and HDL, and acceptability of amaranth plus flaxseed food are the outcome variables of this study.

### Statistical data analysis

The statistical software SPSS version 27 and STATA version16 will be used for data analysis. To determine if the data are normally distributed, a one-sample Kolmogorov–Smirnov test will be used. Mean and standard deviation will be reported for normally distributed data, while median and interquartile ranges will be reported for non-normally distributed data. Parametric analysis will be used to analyse data that will be normally distributed and nonparametric analysis will be used to analyse data that will not be normally distributed. One-way ANOVA will be used to determine the significance of mean difference of scores for colour, appearance, flavour, taste, consistency, mouth fell and overall acceptability (work package I), and also to compare the serum levels of trace elements among the participants (work package II and III). The mean score of proximate composition, mineral, and phytate will analysed by using mean difference to determine the significance change. Each determination of nutrient measurement will carried out on three separate samples and will be analysed in triplicates (work package I). Using the WHO Anthro Plus version 10.4 software, the z score values for height, weight, and BMI for age will be determined (work package II and III). To assess the effects of energy and nutrient-dense amaranth-based food on nutritional status and educational attainments, intention-to-treat (ITT) analysis will be used (work package III). Data will be presented as frequency, percentages, mean, standard deviation, and mean difference. A p-value of <0.05 will be statistically significant in all analyses (work package I-III).

**Figure 2.**
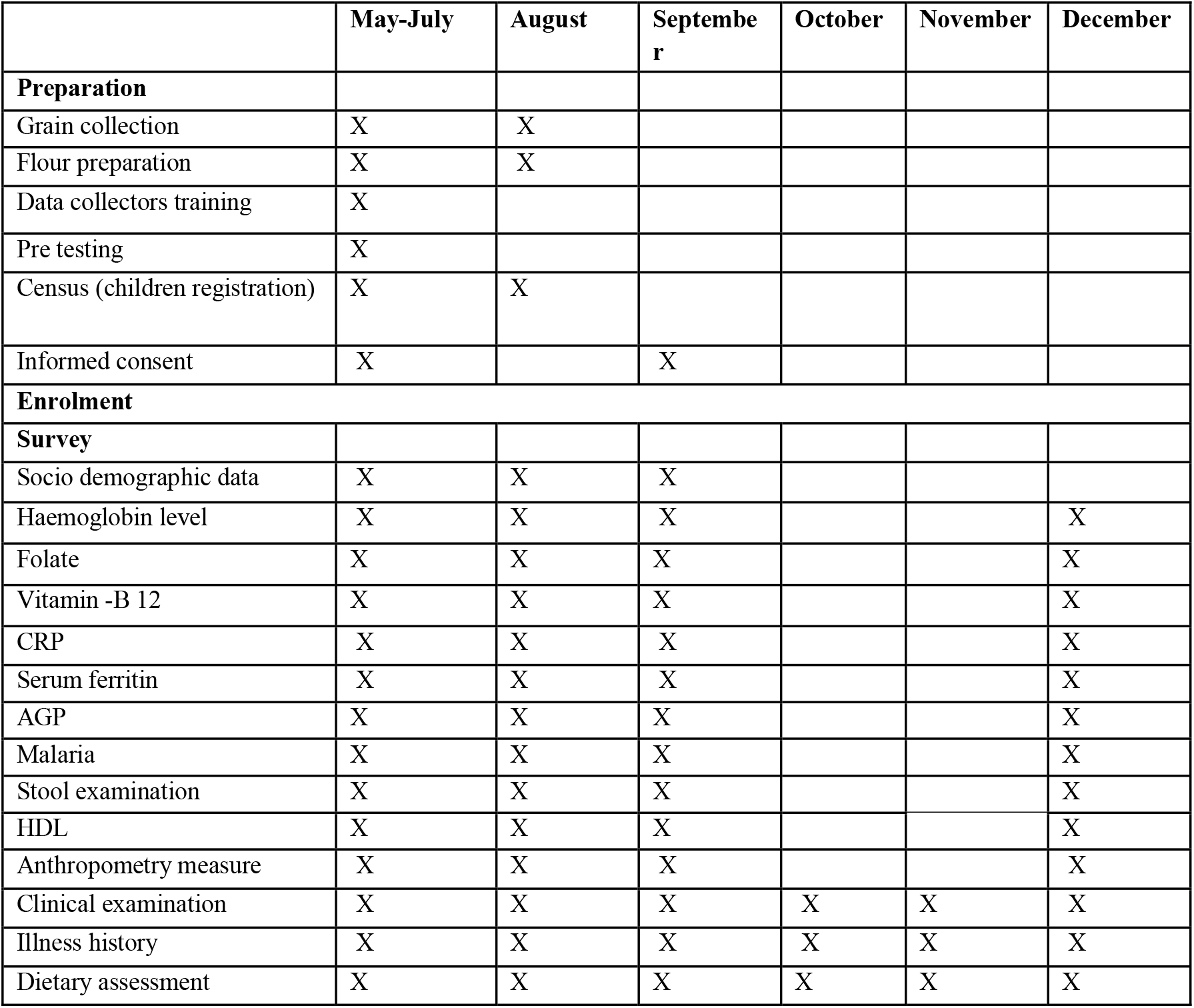
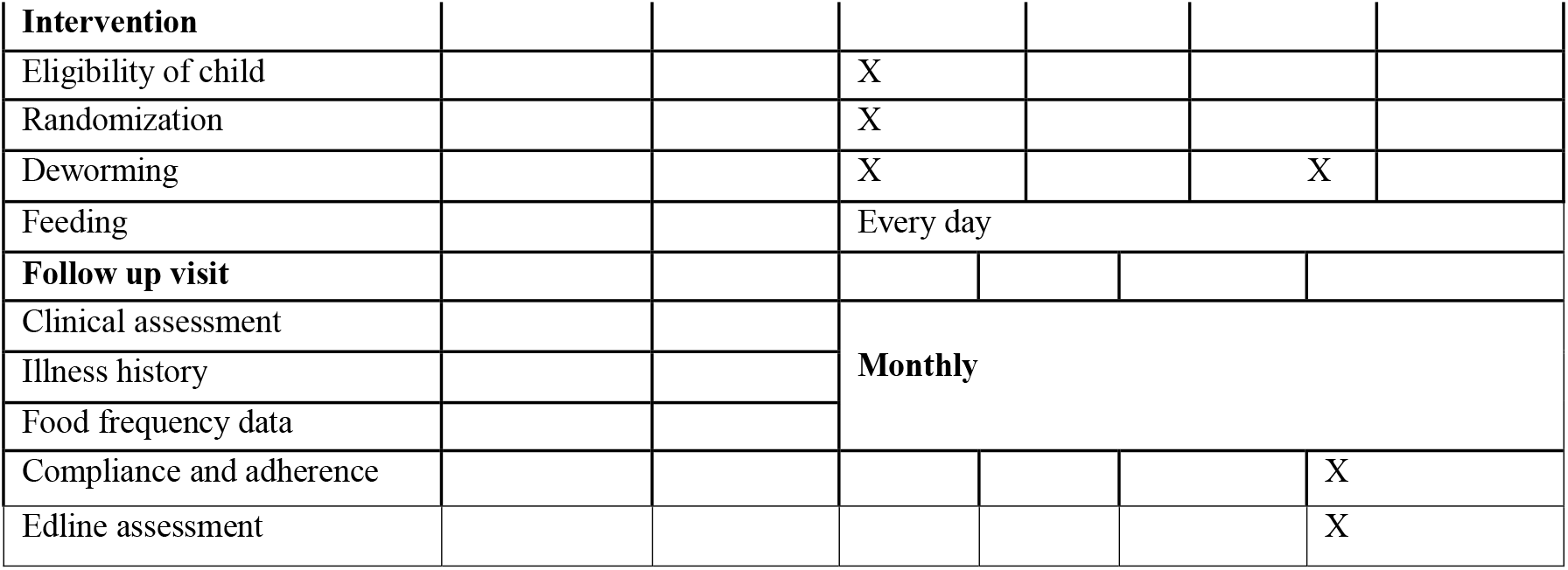
Schedule of enrolment, intervention and assessment of school children, Sidama, Ethiopia, 2024.

### Ethical Considerations

This study protocol has gotten an ethical approval from three institutions including the Institutional Review Board of Hawassa University at the College of Medicine and Health Sciences (Ref.No: IRB/379/15) and Arbaminch University Ethiopia (Ref.No:IRB/23130/2024). As this project is a NORAD-funded project with Norwegian partners, ethical clearance from REK West, Norway. (Ref.No:656883) was sought. Also amendment granted from Hawassa Institutional Review Board of Hawassa University and REK West, Norway. The trial also registered online Pan African Clinical Trial Registry (PACTRA) No: PACTR202403614331028.

Voluntary, written and singed consent will be obtained from each participant from parent and child. All study information will be confidential. To ensure participant privacy, subjects will be assigned unique codes upon enrollment. These codes will be used in all study materials and data analysis instead of names or other identifying information. Only the principal investigator and study staff will have access to the key linking codes to participant identities. All data will be de-identified and stored securely for five years after the study is completed. Study tools will be designed in the local language to facilitate participant understanding.

### Individual advantage and disadvantage from the research

This research aims to benefit all participants. The amaranth/flaxseed group will receive a nutrient-rich diet, while the maize group will benefit from increased calorie intake. The study offers a chance to identify and potentially treat underweight and anemia in children. Participants with severe conditions will be referred to a hospital for further care.

All children will receive anemia treatment based on their needs at the study’s conclusion. Regular monitoring will identify and address any severe nutritional problems, with hospital referrals if needed. Monthly health education sessions will promote healthy habits and feeding practices. While some discomfort might occur during sample collection, standard precautions will be taken to ensure safety.

### Advantage and disadvantage of the research for the community and society

This research holds no foreseeable disadvantages for the community or society. It will benefit the community by highlighting the potential of amaranth and flaxseed as a nutritious food source, potentially increasing farmer income and improving household food security. The findings may also influence policymakers to consider amaranth and flaxseed as alternative food options for school feeding programs, particularly in drought-prone areas. The research will contribute to scientific knowledge regarding the benefits of amaranth and flaxseed for preventing undernutrition. Overall, the potential benefits of this research outweigh the minimal risks involved. The findings will be published openly to support policy decisions and further scientific inquiry.

## Discussion

School feeding programs are one of the ways to mitigate under-nutrition in school children. In spite of its importance, school feeding programs in rural parts of the country are not effective to decrease the malnutrition of school children [32]. The finding from this study may inform policymakers about the usefulness of amaranth and flax seed to improve the nutritional status of the children and costs and sustainability information for potential scale up.

## Data Availability

This article is a research protocol so that no data is required for publication.

## Confidentiality

Participant information will be stored in a secure location and only accessible to authorized personnel and participant identities will be kept anonymity.

## Auditing

The study welcomes any interested organization, whether it be governmental or non-governmental, to conduct an audit.

## Amendment

Regarding protocol modifications, communication will be made to relevant stakeholders, including the IRB/IEC and sponsor.

## Dissemination

The result of this study will be disseminated through Internal Seminars, Conference presentations using outreach and public engagement events. Regular reporting and communication to stakeholders will be done. It will also disseminate through Publications

## Data monitoring commute

This study protocol will be held to test edible food on school feeding therefore there is no anticipated reaction for this preparation.

## Abbreviation

BMI: Body Mass Index
EDHS: Ethiopian Demographic Health Survey
HAZ: Height for Age Z-score
HDL: High Density Lipoprotein
HGSF: Home-Grown School Feeding
FRESH: Focusing Resources on Effective School Health
IDA: Iron Deficiency Anaemia
ICC: Intra Class Correlation Coefficients
MUAC: Mid Upper Arm Circumference
RDI: Recommended Dietary Intake
SHN: School Health Nutrition
USDA: United States Department of Agriculture
WAZ: Weight for Age Z-score
WFP: World Food Program

## Acknowledgment

We would like to acknowledge the government of Norway for funding this study through the Norwegian Programme for Capacity Development in Higher Education and Research for Development (NORHED) providing funding for the South Ethiopia Network Universities in Public Health (SENUPH-II) project

## Trial status

The trial was registered with the Pan-African Clinical Trial Registry on 28 March 2024 and the trial number is PACTR202403614331028.

